# Clinical Characteristics and Outcomes of Laboratory-Confirmed SARS-CoV-2 Cases Infected with Omicron subvariants and XBB recombinant variant

**DOI:** 10.1101/2023.01.05.23284211

**Authors:** Rajesh Karyakarte, Rashmita Das, Sonali Dudhate, Jeanne Agarasen, Praveena Pillai, Priyanka Chandankhede, Rutika Labshetwar, Yogita Gadiyal, Mansi Rajmane, Preeti Kulkarni, Safanah Nizarudeen, Suvarna Joshi, Varsha Potdar, Krishanpal Karmodiya

## Abstract

**Background:** SARS-CoV-2 has evolved to produce new variants causing successive waves of infection. Currently, six variants are being monitored by the World Health Organization that are replacing BA.5. These include BF.7 (BA.5 + R346T in spike), BQ.1 (and BQ.1.1, with BA.5 + R346T, K444T, N460K mutations in spike), BA.2.75 (including BA.2.75.2 and CH.1.1), and XBB (including XBB.1.5). BQ.1 and XBB variants are more immune evasive and have spread quickly throughout the world. With the concern of the potential severity of infections caused by these variants, the present study describes the clinical characteristics and outcomes of these major variants in Maharashtra.

**Material and Methods:** A total of 1141 Reverse Transcriptase-Polymerase Chain Reaction (RT-PCR) positive SARS-CoV-2 samples, with a cycle threshold value (Ct) less than 25, were processed for SARS-CoV-2 whole genome sequencing between 10th July 2022 and 12th January 2023. All corresponding demographic and clinical data were recorded and analysed using Microsoft® Excel and Epi Info™.

**Results:** Out of 1141 samples sequenced, BA.2.75* (63.78%) was the predominant Omicron variant, followed by the XBB* (18.88%), BA.2.38* (4.94%), BA.5* (4.06%), BA.2.10* (3.51%) and BQ.1* (1.65%). A total of 540 cases were contacted telephonically, of which 494 (91.48%) were symptomatic with mild symptoms. Fever (77.73%) was the most common symptom, followed by cold (47.98%), cough (42.31%), myalgia and fatigue (18.83%). Of the 540 cases, 414 (76.67%) cases recovered at home, and 126 (23.33%) were institutionally quarantined/hospitalised. Among the home-isolated and hospitalised cases, 416 (99.76%) and 108 (87.80%), respectively, recovered with symptomatic treatment, while one (0.24%) and 15 (12.20%), respectively, succumbed to the disease. In all, 491 (90.93%) cases were vaccinated with at least one dose of the COVID-19 vaccine, 41 (7.59%) were unvaccinated, and for 08 (1.48%), vaccine data was not available.

**Conclusion:** The current study indicates that the XBB* variant is causing mild disease in India. However, as XBB* possess both immune-escape and infectivity-enhancing mutations, it has the potential to spread to other parts of the world rapidly.

## 1. INTRODUCTION

The Severe Acute Respiratory Syndrome Coronavirus 2 (SARS-CoV-2) has evolved continuously to give rise to new variants driving successive waves of infection. (1) Since its first emergence in November 2021, the Omicron variant of concern (VOC) has become the most widespread and dominant variant globally. Currently, BA.5 and its descendant lineages are the dominant variants, followed by BA.2.75 and its subvariants (as of 1^st^ January 2023). (2) With a swarm of variants emerging and competing in the “variant soup” (3), there are four subvariants which are replacing the BA.5 descendent lineages and are under monitoring by the World Health Organization. (2) These include BF.7 (BA.5 + R346T in spike), BQ.1 (and BQ.1.1, with BA.5 + R346T, K444T, N460K mutations in spike), BA.2.75 (including BA.2.75.2 and CH.1.1), and XBB (including XBB.1.5). (2) BQ.1, a sub-lineage of BA.5, was first identified in Nigeria in early July 2022. There are 80 Pango lineages associated with the BQ.1* variant (* indicates the lineage and its sub-lineages), which have spread dramatically to Europe, South America, North America, and Africa. (4,5) XBB is a recombinant of BA.2.10.1 and BA.2.75 sub-lineages, i.e., BJ.1 and BM.1.1.1, respectively, with a breakpoint in S1. It was first identified in mid-August 2022 and is suggested to have emerged around the Indian subcontinent. (5) XBB and its sub-lineages have spread quickly and have become dominant in India, Bangladesh, Malaysia, Singapore, and other parts of Asia. (5,6) Recently, a subvariant of XBB.1, the XBB.1.5 variant, has rapidly spread and has been detected in at least 46 countries and 49 states in the United States of America. (7) As these variants continue to evolve and diversify, they are of particular interest, as they are more immune evasive and expand rapidly due to additional mutations in their spike protein. There is evidence that these SARS-CoV-2 variants may further reduce the effectiveness of current COVID-19 vaccines and monoclonal antibody treatments. (5)

For an effective pandemic response, it is essential to understand the range of illnesses associated with these new strains. There are concerns about the potential severity of infections caused by these variants due to multiple mutations in their spike protein, which may affect their ability to enter cells and escape the immune system. (8) Therefore, the present study describes the clinical characteristics and outcomes of the major variants identified during the community surveillance of SARS-CoV-2 in Maharashtra.

## 2. MATERIAL AND METHODS

The present study was conducted as part of the Indian SARS-CoV-2 Genomics Consortium (INSACOG) sequencing activity in Maharashtra to monitor genomic variations in the virus and to study its epidemiological trends. The study protocol for SARS-CoV-2 whole genome sequencing was reviewed and approved by the Institutional Ethics Committee at Byramjee Jeejeebhoy Government Medical College (BJGMC), Pune, Indian Council of Medical Research-National Institute of Virology (ICMR-NIV), Pune and Indian Institute of Science, Education and Research (IISER), Pune.

### 2.1 Sample Acquisition

Samples from several RT-PCR swab collection and testing centres in various districts of Maharashtra and its neighbouring states were sent to BJGMC, ICMR-NIV and IISER, Pune, for whole genome sequencing.

Respiratory specimens, including nasopharyngeal and oropharyngeal swabs, positive for SARS-CoV-2 infection, were collected in Viral Transport Medium (VTM). These samples were transported to the sequencing laboratory in triple packaging, maintaining the cold chain, and stored at -80°C. According to government instructions and INSACOG, 5% of the samples from positive cases with a Ct value less than 25, including clusters, vaccine breakthrough infections, cases with mild and moderate symptoms, and hospitalised and deceased cases, were processed.

### 2.2 RNA Extraction, Library Preparation, Next Generation Sequencing, and Lineage Analysis

Total RNA was extracted from respiratory specimens using the MagMax™ Viral/Pathogen nucleic acid extraction kit (ThermoFisher Scientific, Waltham, MA, USA) on an automated extraction system, KingFisher Flex (ThermoFisher, Waltham, MA, USA), following the manufacturer’s instructions. Nucleic acid was eluted in 50 μL of elution buffer, and the RNA was quantified with the Qubit RNA High Sensitivity Kit using the Qubit® 2.0 Fluorometer (ThermoFisher Scientific, Waltham, MA, USA). This RNA was used for library preparation for sequencing.

Libraries for SARS-CoV-2 sequencing were prepared using the Rapid Barcoding Kit (RBK110.96) and Midnight RT-PCR Expansion Kits (EXP001) (BJGMC, Pune), Ion 540TM chip and the Ion Total RNA-Seq kit v2.0 (ThermoFisher Scientific, Waltham, MA, USA) (ICMR-NIV, Pune) and Illumina COVIDSeq RUO test kits (Illumina Inc, USA) (IISER, Pune). Sequencing was performed using GridION (ONT, Littlemore, United Kingdom), Ion S5 (ThermoFisher Scientific, Waltham, MA, USA) and the NextSeq 550 sequencing platform, respectively. Reads were aligned to the reference genome using MinKNOW software, Iterative Refinement Meta-Assembler (IRMA) and DRAGEN COVID Lineage application, respectively. Lineage analysis was done using Phylogenetic Assignment of Named Global Outbreak LINeages (PangoLIN) COVID-19 lineage assigner, version v4.1.3, pangolin-data version v1.17, and clade was analysed using Nextclade software, version v2.9.1.

### 2.3 Demographic and Clinical Data Collection

A set of demographic data, including the patient’s age, gender, area of residence, contact number, and date of specimen collection and testing, were collected from the centralised data entry portal for COVID-19 testing in India, the ICMR COVID-19 Data Portal using the unique identification number (ICMR ID). In order to gather additional information, telephonic interviews were conducted, and patients were interviewed individually. During the interview, the demographic details available on the portal were validated, and information on the presence of any symptoms at the time of testing, comorbidities, vaccination details, history of the previous infection, hospitalisation, oxygen requirement and treatment were collected. Patients unwilling to share their history during the interview were excluded from the study.

### 2.4 Statistical Analysis

All demographic and clinical data were recorded using Microsoft® Excel, and analysis was performed using Microsoft® Excel and Epi Info™, version 7.2.4.0. The continuous variables were presented as the median and interquartile range (IQR). The Kruskal-Wallis test was performed to compare the median values between the variants. The categorical variables were presented as numbers and percentages. The Chi-square test was used to compare categorical variables between the variants. Fisher’s exact test compared the categorical values with limited data. A *p*-value < 0.05 was considered statistically significant.

## 3. RESULTS

### 3.1 Demographic Characteristics of the Study Population

Between 10th July 2022 and 12th January 2023, 1141 RT-PCR-positive SARS-CoV-2 samples were collected and included in the study. ***Table 1*** describes the geographical distribution of the samples collected. The study population included cases from all age groups with a median age of 37 years (IQR: 28-57) (***Table 2***). The male-to-female ratio was 1.22:1 (***Table 3***).

**Table 1:** Geographical distribution of 1141 RT-PCR-positive SARS-CoV-2 samples

| Area of Residence |  | Frequency | Percentage | 95% LCL | 95% UCL |
| --- | --- | --- | --- | --- | --- |
| Maharashtra | Ahmednagar | 7 | 0.61% | 0.30% | 1.26% |
|  | Akola | 4 | 0.35% | 0.14% | 0.90% |
|  | Alibag | 17 | 1.49% | 0.93% | 2.37% |
|  | Amravati | 5 | 0.44% | 0.19% | 1.02% |
|  | Aurangabad | 4 | 0.35% | 0.14% | 0.90% |
|  | Beed | 11 | 0.96% | 0.54% | 1.72% |
|  | Buldhana | 2 | 0.18% | 0.05% | 0.64% |
|  | Jalgaon | 2 | 0.18% | 0.05% | 0.64% |
|  | Jalna | 7 | 0.61% | 0.30% | 1.26% |
|  | Kolhapur | 94 | 8.24% | 6.78% | 9.98% |
|  | Mumbai | 76 | 6.66% | 5.35% | 8.26% |
|  | Nagpur | 1 | 0.09% | 0.02% | 0.49% |
|  | Nanded | 1 | 0.09% | 0.02% | 0.49% |
|  | Nashik | 2 | 0.18% | 0.05% | 0.64% |
|  | Osmanabad | 2 | 0.18% | 0.05% | 0.64% |
|  | Pune | 801 | 70.20% | 67.48% | 72.78% |
|  | Satara | 13 | 1.14% | 0.67% | 1.94% |
|  | Solapur | 1 | 0.09% | 0.02% | 0.49% |
|  | Thane | 73 | 6.40% | 5.12% | 7.97% |
|  | Washim | 1 | 0% | 0.02% | 0.49% |
| Outside Maharashtra | Goa | 15 | 1.31% | 0.80% | 2.16% |
|  | Silavasa | 2 | 0.18% | 0.05% | 0.64% |
| TOTAL |  | 1141 | 100% |  |  |

**Table 2:**
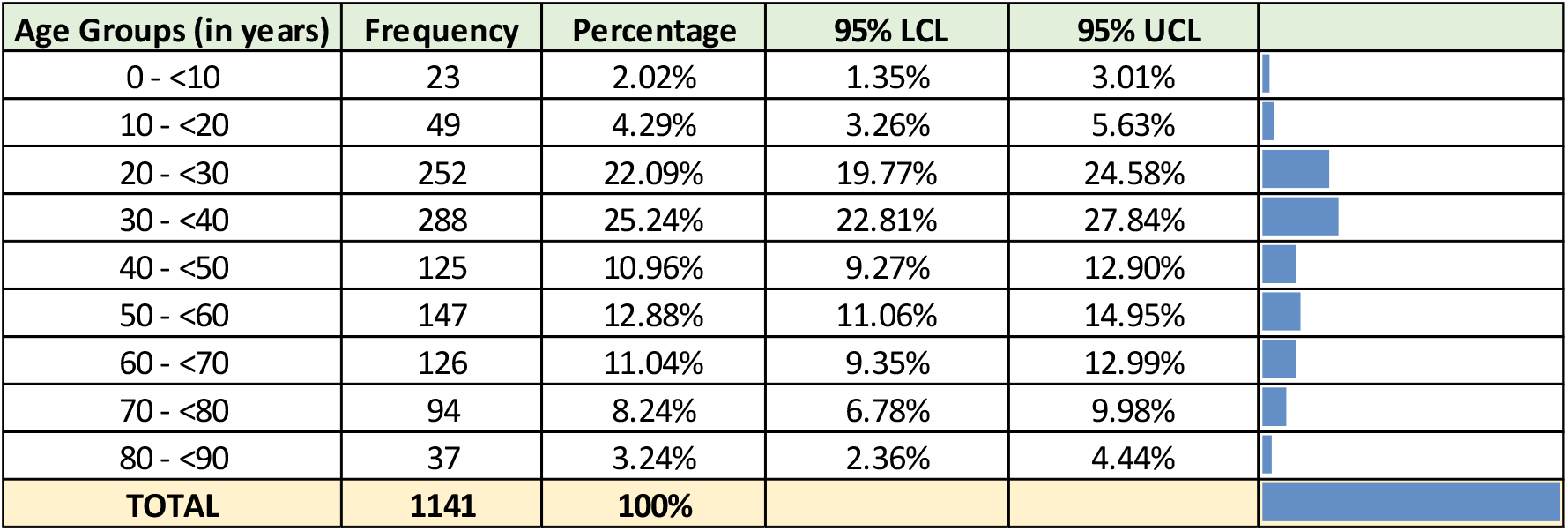
Age-wise distribution of 1141 RT-PCR-positive SARS-CoV-2 samples

| Age Groups (in years) | Frequency | Percentage | 95% LCL | 95% UCL |
| --- | --- | --- | --- | --- |
| 0 - <10 | 23 | 2.02% | 1.35% | 3.01% |
| 10 - <20 | 49 | 4.29% | 3.26% | 5.63% |
| 20 - <30 | 252 | 22.09% | 19.77% | 24.58% |
| 30 - <40 | 288 | 25.24% | 22.81% | 27.84% |
| 40 - <50 | 125 | 10.96% | 9.27% | 12.90% |
| 50 - <60 | 147 | 12.88% | 11.06% | 14.95% |
| 60 - <70 | 126 | 11.04% | 9.35% | 12.99% |
| 70 - <80 | 94 | 8.24% | 6.78% | 9.98% |
| 80 - <90 | 37 | 3.24% | 2.36% | 4.44% |
| TOTAL | 1141 | 100% |  |  |

**Table 3:**
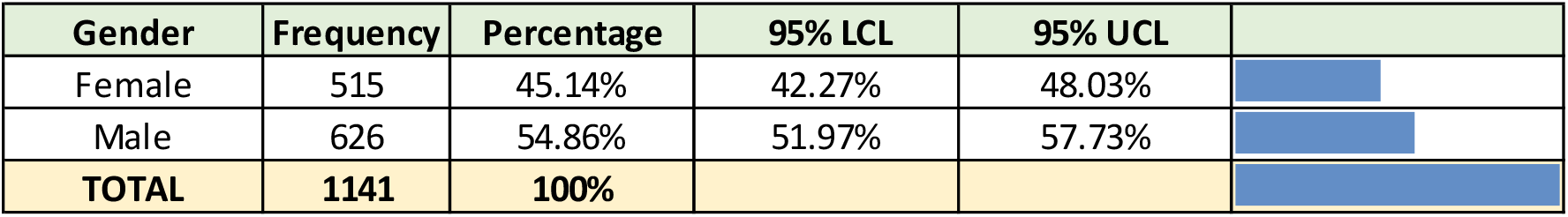
Gender-wise distribution of 1141 RT-PCR-positive SARS-CoV-2 samples

| Gender | Frequency | Percentage | 95% LCL | 95% UCL |
| --- | --- | --- | --- | --- |
| Female | 515 | 45.14% | 42.27% | 48.03% |
| Male | 626 | 54.86% | 51.97% | 57.73% |
| **TOTAL** | **1141** | **100%** |  |  |

### 3.2 SARS-CoV-2 Lineage Distribution in Sequenced Samples

Of the 1141 samples sequenced, 911 (79.84%) were assigned Pango lineages, of which the BA.2.75* (63.78%) was the predominant Omicron variant, followed by the XBB* (18.88%), BA.2.38* (4.94%), BA.5* (4.06%), BA.2.10* (3.51%) and BQ.1* (1.65%) (***Table 4***). ***Figure 1*** describes the temporal distribution of SARS-CoV-2 variants detected during the study period. The x-axis represents the calendar weeks, and the y-axis represents the percentage of each lineage from the sequenced samples.

**Table 4:**
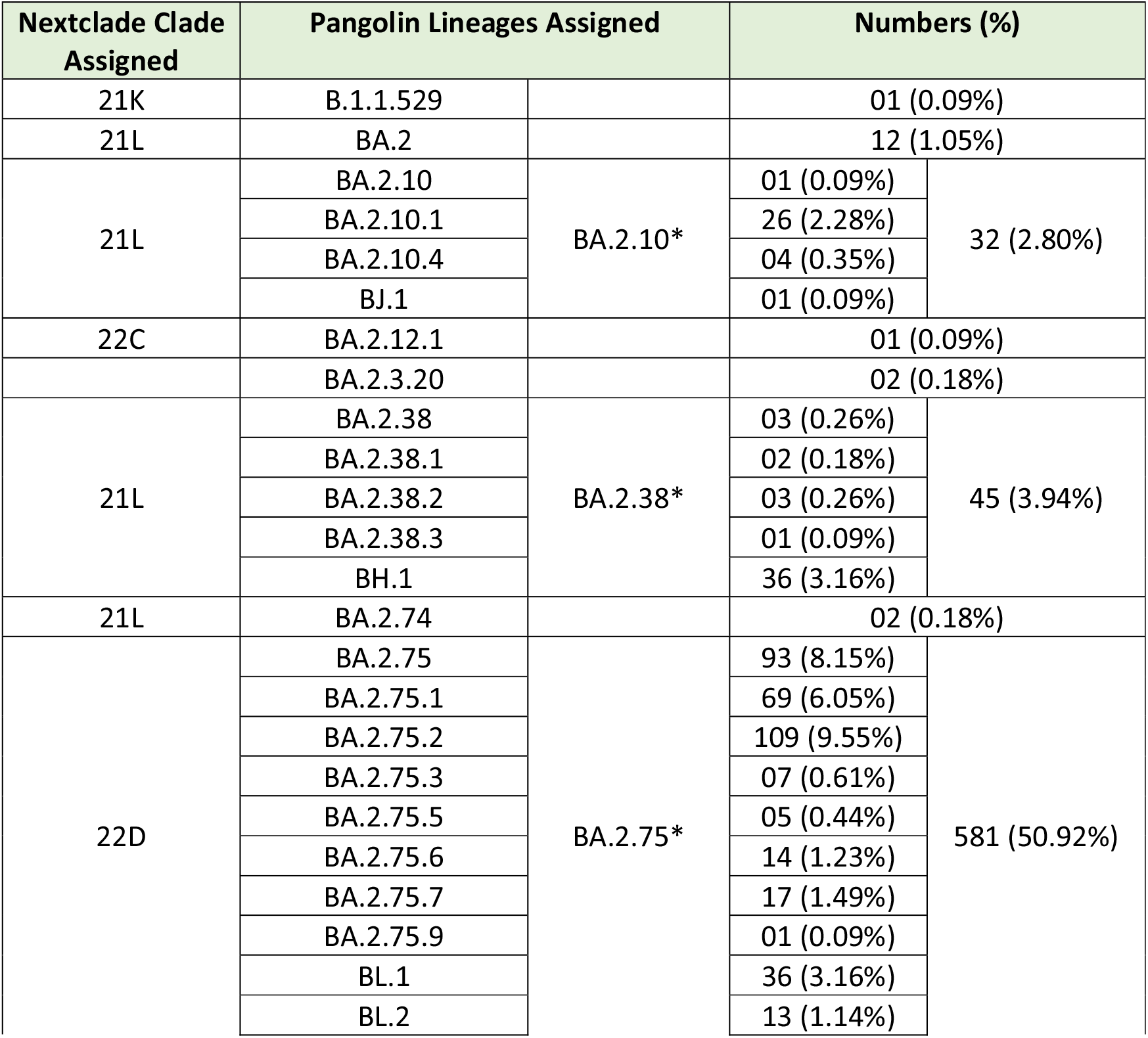
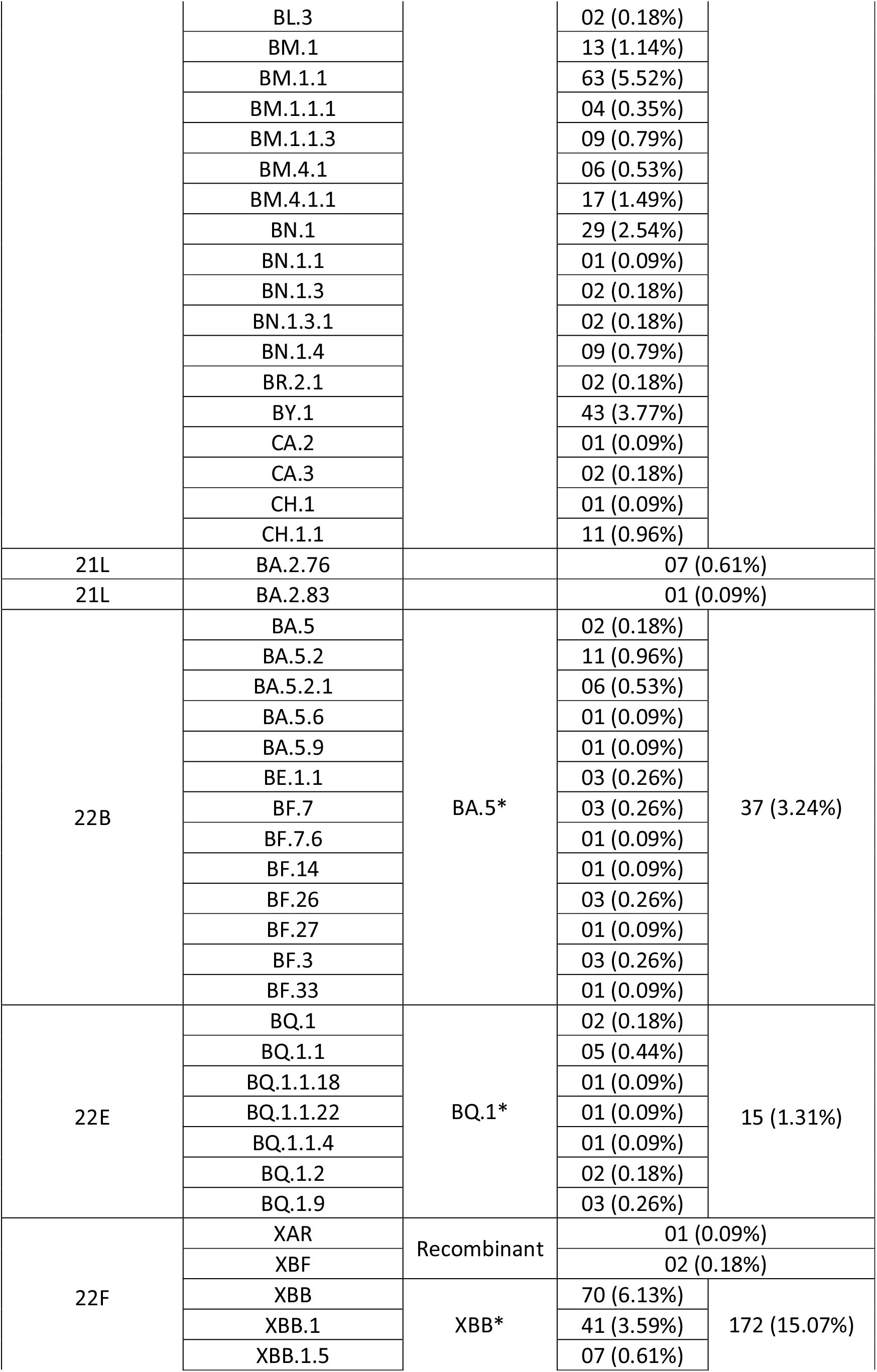
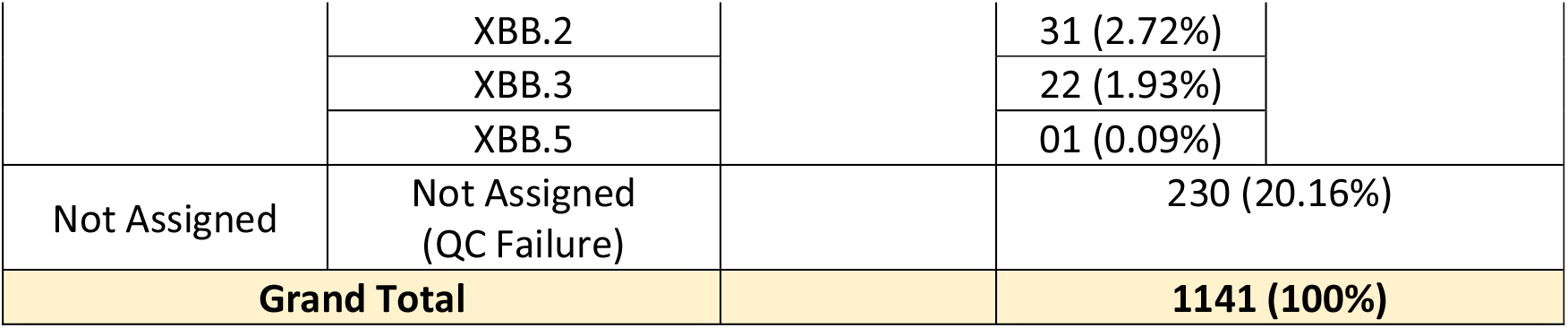
Variant distribution among the 1141 SARS-CoV-2 samples sequenced

**Figure 1:**
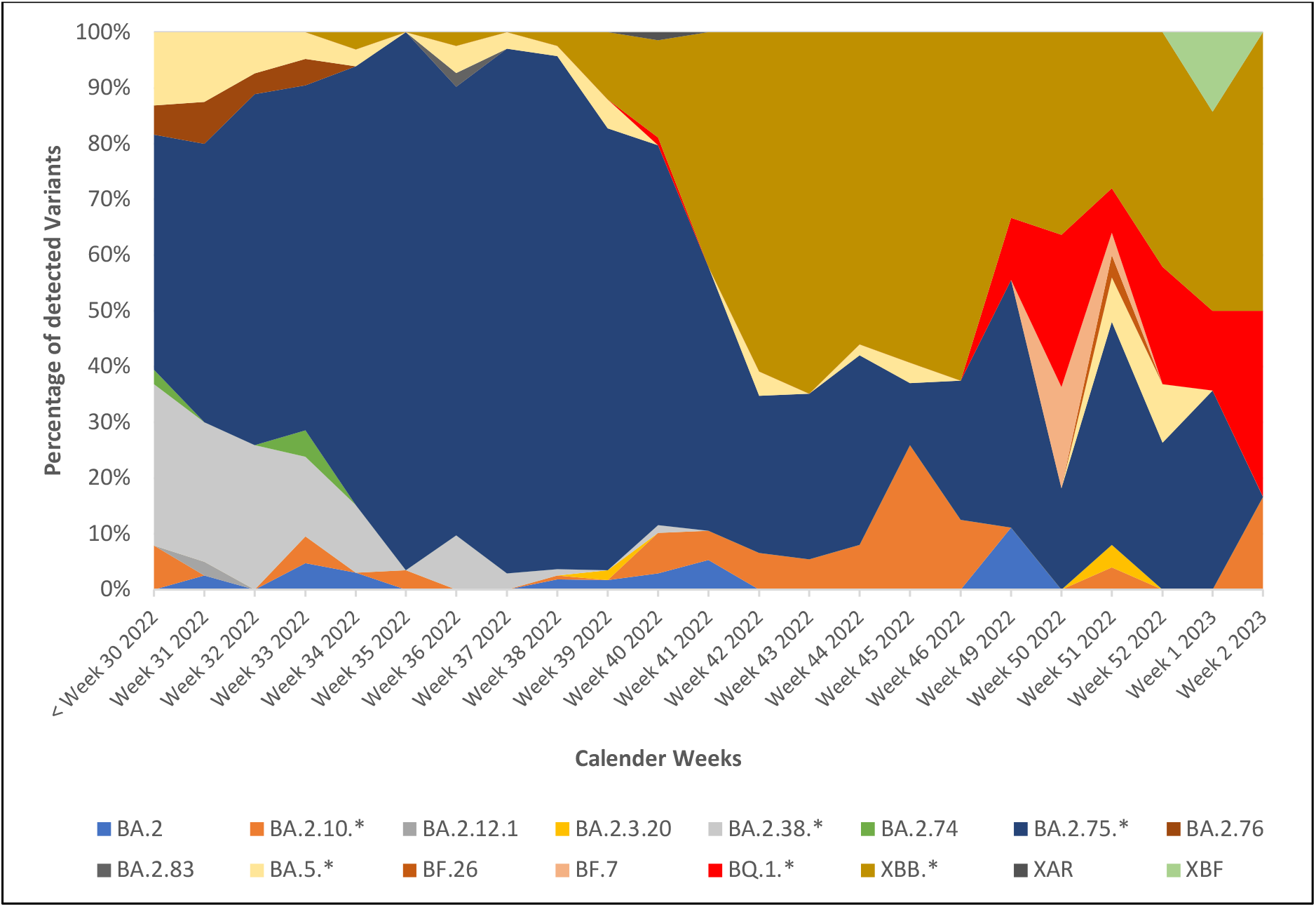
Temporal distribution of SARS-CoV-2 variants during the study period (From 10^th^ July 2022 to 12^th^ January 2023)

### 3.3 Demographic and clinical characteristics of major SARS-CoV-2 variants detected during the study

***Table 5*** describes the demographic characteristics of the major SARS-CoV-2 variants detected in the study. Out of 882 cases, there were 475 (53.85%) males and 407 (46.15%) females. The median age of cases infected with BA.2.10* was 29.5 years, BQ.1* was 55.0 years, XBB* was 38.5 years, and for BA.2.38*, BA.2.75*, BA.5* variants, it was 38.0 years. There was a statistically significant difference in the median age (*p*-value = 0.030) of cases infected with different variants. ***Figure 2*** depicts the distribution of major variants and the area of residence of cases.

**Table 5:** Demographic characteristics of major SARS-CoV-2 variants detected during the study period (n=882)

|  | BA.2.10* | BA.2.38* | BA.2.75* | BA.5* | BQ.1* | XBB* | Grand Total |
| --- | --- | --- | --- | --- | --- | --- | --- |
| **1. Gender-wise distribution** |  |  |  |  |  |  | **p - value = 0.088** |
| **Male** | 22<br>(68.75%) | 19<br>(42.22%) | 311<br>(53.53%) | 15<br>(40.54%) | 10<br>(66.67%) | 98<br>(56.98%) | 475<br>(53.85%) |
| **Female** | 10<br>(31.25%) | 26<br>(57.78%) | 270<br>(46.47%) | 22<br>(59.46%) | 05<br>(33.33%) | 74<br>(43.02%) | 407<br>(46.15%) |
| **2. Median Age** |  |  |  |  |  |  | **H (5) = 12.343, p-value = 0.030** |
|  | 29.5<br>(IQR: 20.5 – 43.0) | 38.0<br>(IQR: 29.0 – 58.0) | 38.0<br>(IQR: 28.0 – 57.0) | 38.0<br>(IQR: 31.0 – 59.0) | 55.0<br>(IQR: 41.0 – 71.0) | 38.5<br>(IQR: 28.5 – 59.0) |  |
| **3. Age-wise distribution** |  |  |  |  |  |  |  |
| **0 - <10** | 01<br>(3.13%) | 00<br>(00%) | 11<br>(1.89%) | 01<br>(2.70%) | 00<br>(00%) | 03<br>(1.74%) | 16<br>(1.81%) |
| **10 - <20** | 07<br>(21.88%) | 04<br>(8.89%) | 25<br>(4.30%) | 00<br>(00%) | 00<br>(00%) | 11<br>(6.40%) | 47<br>(5.33%) |
| **20 - <30** | 08<br>(25.00%) | 08<br>(17.78%) | 133<br>(22.89%) | 06<br>(16.22%) | 01<br>(6.67%) | 32<br>(18.60%) | 188<br>(21.32%) |
| **30 - <40** | 04<br>(12.50%) | 11<br>(24.44%) | 140<br>(24.10%) | 13<br>(35.14%) | 02<br>(13.33%) | 42<br>(24.42%) | 212<br>(24.04%) |
| **40 - <50** | 05<br>(15.63%) | 06<br>(13.33%) | 53<br>(9.12%) | 03<br>(8.11%) | 04<br>(26.67%) | 21<br>(12.21%) | 92<br>(10.43%) |
| **50 - <60** | 01<br>(3.13%) | 05<br>(11.11%) | 86<br>(14.80%) | 07<br>(18.92%) | 02<br>(13.33%) | 22<br>(12.79%) | 123<br>(13.95%) |
| **60 - <70** | 05<br>(15.63%) | 04<br>(8.89%) | 64<br>(11.02%) | 05<br>(13.51%) | 02<br>(13.33%) | 18<br>(10.47%) | 98<br>(11.11%) |
| **70 - <80** | 01<br>(3.13%) | 06<br>(13.33%) | 48<br>(8.26%) | 02<br>(5.41%) | 03<br>(20.00%) | 18<br>(10.47%) | 78<br>(8.84%) |
| **80 - <90** | 00<br>(00%) | 01<br>(2.22%) | 21<br>(3.61%) | 00<br>(00%) | 01<br>(6.67%) | 05<br>(2.91%) | 28<br>(3.17%) |

**Figure 2:**
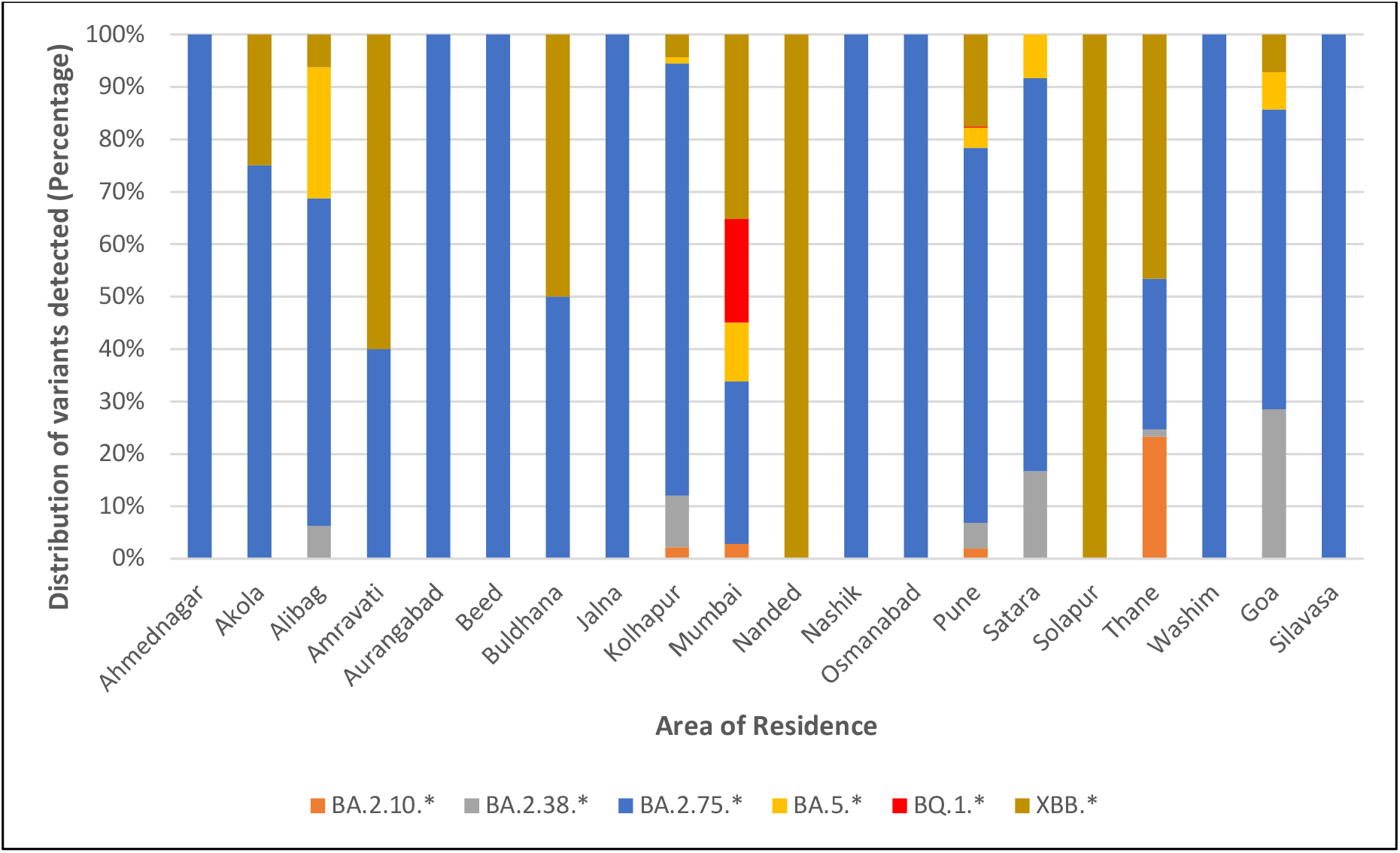
Distribution of major SARS-CoV-2 variants detected during the study versus the area of residence

Of the 882 cases, 540 (61.22%) could be telephonically contacted to obtain information regarding their symptoms, hospitalisation, treatment and vaccination status. ***Table 6*** summarises the clinical characteristics, vaccination status and outcome of 494 cases. Most cases were symptomatic (91.48%) with mild symptoms. Fever (77.73%) was the most common symptom across all variants, followed by cold and rhinorrhoea (47.98%), cough (42.31%), myalgia and fatigue (18.83%). Of the 540 cases, 478 (88.52%) cases confirmed the presence of no comorbidity. Among those with one or more comorbid conditions, diabetes mellitus was the most common condition reported (51.61%), followed by hypertension (50%), carcinoma (8.06%), coronary heart disease (6.45%), liver and kidney disease (4.84%), asthma (4.84%) tuberculosis and arthritis (3.23% each). There was a statistically significant difference between the absence or presence of comorbidity and the outcome of disease (*p*-value=<0.001) (***Figure 3***). There were 414 (76.67%) cases who required home isolation, and 126 (23.33%) cases were institutionally quarantined/hospitalised. Of the 540 cases, 3.15% recovered without any treatment, 87.96% recovered with supportive treatment, 6.67% required oxygen therapy and 2.22% were given antiviral treatment. There were 16 (2.96%) cases who succumbed to the disease, and the rest (97.04%) survived. ***Figure 4*** depicts the age-wise distribution of survived and dead cases.

**Table 6:** Clinical Characteristics and outcome of 540 patients infected with major variants (n=540)

|  | BA.2.10* | BA.2.38* | BA.2.75* | BA.5* | BQ.1 | XBB* | Grand Total |
| --- | --- | --- | --- | --- | --- | --- | --- |
| **Total number of cases with history available** |  |  |  |  |  |  |  |
|  | 17<br>(53.13%) | 21<br>(46.67%) | 374<br>(64.37%) | 18<br>(48.65%) | 10<br>(66.67%) | 100<br>(58.14%) | 540<br>(61.22%) |
| **History of previous COVID-19 infection** **p - value = 0.259** |  |  |  |  |  |  |  |
|  | 01<br>(5.88%) | 03<br>(14.29%) | 51<br>(13.64%) | 00<br>(00%) | 01<br>(10%) | 15<br>(15%) | 71<br>(13.15%) |
| **Symptom status at the time of sample collection** **p - value = 0.071** |  |  |  |  |  |  |  |
| Asymptomatic | 03<br>(17.65%) | 01<br>(4.76%) | 28<br>(7.49%) | 00<br>(00%) | 03<br>(30%) | 11<br>(11%) | 46<br>(8.52%) |
| Symptomatic | 14<br>(82.35%) | 20<br>(95.24%) | 346<br>(92.51%) | 18<br>(100%) | 07<br>(70%) | 89<br>(89%) | 494<br>(91.48%) |
| **Presence of comorbidity** **p - value = 0.145** |  |  |  |  |  |  |  |
| No comorbidity | 17<br>(100%) | 15<br>(71.43%) | 334<br>(89.30%) | 17<br>(94.44%) | 10<br>(100%) | 85<br>(85%) | 478<br>(88.52%) |
| Presence of one condition | 00<br>(00%) | 05<br>(23.81%) | 27<br>(7.22%) | 01<br>(5.56%) | 00<br>(00%) | 11<br>(11%) | 44<br>(8.15%) |
| Presence of two or more conditions | 00<br>(00%) | 01<br>(4.76%) | 13<br>(3.48%) | 00<br>(00%) | 00<br>(00%) | 04<br>(4%) | 18<br>(3.33%) |
| **Initial presenting symptoms** |  |  |  |  |  |  |  |
| Fever | 08<br>(57.14%) | 16<br>(80%) | 277<br>(80.06%) | 13<br>(72.22%) | 05<br>(71.43%) | 65<br>(73.03%) | 384<br>(77.73%) |
| Cough | 05<br>(35.71%) | 13<br>(65%) | 146<br>(42.20%) | 07<br>(38.89%) | 04<br>(57.14%) | 34<br>(38.20%) | 209<br>(42.31%) |
| Fatigue/ Weakness | 00<br>(00%) | 02<br>(10%) | 78<br>(22.54%) | 02<br>(11.11%) | 01<br>(14.29%) | 10<br>(11.24%) | 93<br>(18.83%) |
| Myalgia | 03<br>(21.43%) | 00<br>(00%) | 74<br>(21.39%) | 01<br>(5.56%) | 02<br>(28.57%) | 13<br>(14.61%) | 93<br>(18.83%) |
| Cold and Rhinorrhoea | 07<br>(50%) | 06<br>(30%) | 163<br>(47.11%) | 11<br>(61.11%) | 03<br>(42.86%) | 47<br>(52.81%) | 237<br>(47.98%) |
| Headache | 00<br>(00%) | 02<br>(10%) | 26<br>(7.51%) | 00<br>(00%) | 02<br>(28.57%) | 10<br>(11.24%) | 40<br>(8.10%) |
| Diarrhoea | 00<br>(00%) | 00<br>(00%) | 05<br>(1.45%) | 02<br>(11.11%) | 00<br>(00%) | 03<br>(3.37%) | 10<br>(2.02%) |
| Breathlessness | 00<br>(00%) | 02<br>(10%) | 22<br>(6.36%) | 00<br>(00%) | 01<br>(14.29%) | 06<br>(6.74%) | 31<br>(6.28%) |
| Vomiting | 00<br>(00%) | 00<br>(00%) | 09<br>(2.60%) | 00<br>(00%) | 00<br>(00%) | 02<br>(2.25%) | 11<br>(2.42%) |
| Sore throat | 01<br>(7.14%) | 01<br>(5%) | 15<br>(4.34%) | 01<br>(5.56%) | 02<br>(28.57%) | 16<br>(17.98%) | 36<br>(7.29%) |
| Skin rash | 00<br>(00%) | 00<br>(00%) | 03<br>(0.87%) | 00<br>(00%) | 00<br>(00%) | 00<br>(00%) | 03<br>(0.61%) |
| Chest pain | 00<br>(00%) | 00<br>(00%) | 01<br>(0.29%) | 00<br>(00%) | 00<br>(00%) | 00<br>(00%) | 01<br>(0.20%) |
| Loss of taste | 00<br>(00%) | 00<br>(00%) | 02<br>(0.58%) | 00<br>(00%) | 00<br>(00%) | 02<br>(2.25%) | 04<br>(0.81%) |
| Loss of smell | 00<br>(00%) | 00<br>(00%) | 02<br>(0.58%) | 00<br>(00%) | 00<br>(00%) | 01<br>(1.12%) | 03<br>(0.61%) |
| **Type of Quarantine** **p - value = 0.163** |  |  |  |  |  |  |  |
| Home quarantine | 16<br>(94.12%) | 14<br>(66.67%) | 280<br>(74.87%) | 15<br>(83.33%) | 09<br>(90%) | 80<br>(80%) | 414<br>(76.67%) |
| Institutional quarantine/<br>required hospitalisation | 01<br>(5.88%) | 07<br>(33.33%) | 94<br>(25.13%) | 03<br>(16.67%) | 01<br>(10%) | 20<br>(20%) | 126<br>(23.33%) |

| Treatment *p* - value = 0.067 |  |  |  |  |  |  |  |
| --- | --- | --- | --- | --- | --- | --- | --- |
| No treatment taken | 03<br>(17.65%) | 00<br>(00%) | 10<br>(2.67%) | 00<br>(00%) | 00<br>(00%) | 04<br>(4%) | 17<br>(3.15%) |
| Need for conservative treatment | 13<br>(76.47%) | 17<br>(80.95%) | 330<br>(88.24%) | 18<br>(100%) | 10<br>(100%) | 87<br>(87%) | 475<br>(87.96%) |
| Need for supplemental oxygen | 01<br>(5.88%) | 04<br>(19.05%) | 27<br>(7.22%) | 00<br>(00%) | 00<br>(00%) | 05<br>(5%) | 37<br>(6.85%) |
| a. Low flow oxygen | 01<br>(100%) | 03<br>(75%) | 23<br>(85.18%) | 00<br>(00%) | 00<br>(00%) | 04<br>(80%) | 31<br>(83.78%) |
| b. Intubation | 00<br>(00%) | 01<br>(25%) | 04<br>(14.82%) | 00<br>(00%) | 00<br>(00%) | 01<br>(20%) | 06<br>(16.22%) |
| Need for antiviral agents/steroids or immunomodulatory drugs | 00<br>(00%) | 00<br>(00%) | 07<br>(1.87%) | 00<br>(00%) | 00<br>(00%) | 04<br>(4%) | 11<br>(2.04%) |
| The outcome of disease *p* - value = 0.495 |  |  |  |  |  |  |  |
| Survived | 17<br>(100%) | 21<br>(100%) | 362<br>(96.79%) | 18<br>(100%) | 10<br>(100%) | 96<br>(96%) | 524<br>(97.04%) |
| Dead | 00<br>(00%) | 00<br>(00%) | 12<br>(3.21%) | 00<br>(00%) | 00<br>(00%) | 04<br>(4%) | 16<br>(2.96%) |
| Vaccination Status *p* - value = 0.181 |  |  |  |  |  |  |  |
| Vaccinated with at least one dose | 13<br>(76.47%) | 18<br>(85.71%) | 341<br>(91.18%) | 15<br>(83.33%) | 10<br>(100%) | 94<br>(94%) | 491<br>(90.93%) |
| a. Single Dose | 00<br>(00%) | 00<br>(00%) | 15<br>(4.40%) | 00<br>(00%) | 00<br>(00%) | 02<br>(2.13%) | 17<br>(3.46%) |
| b. Two doses | 11<br>(84.62%) | 16<br>(88.89%) | 238<br>(69.79%) | 11<br>(73.33%) | 06<br>(60%) | 68<br>(72.34%) | 329<br>(73.27%) |
| c. Booster Dose | 02<br>(15.38%) | 02<br>(11.11%) | 88<br>(25.81%) | 04<br>(26.67%) | 04<br>(40%) | 24<br>(25.53%) | 104<br>(23.16%) |
| Not vaccinated | 04<br>(23.53%) | 03<br>(14.29%) | 28<br>(7.49%) | 03<br>(16.67%) | 00<br>(00%) | 03<br>(3%) | 41<br>(7.59%) |
| Data not available | 00<br>(00%) | 00<br>(00%) | 05<br>(1.34%) | 00<br>(00%) | 00<br>(00%) | 03<br>(3%) | 08<br>(1.48%) |

**Figure 3:**
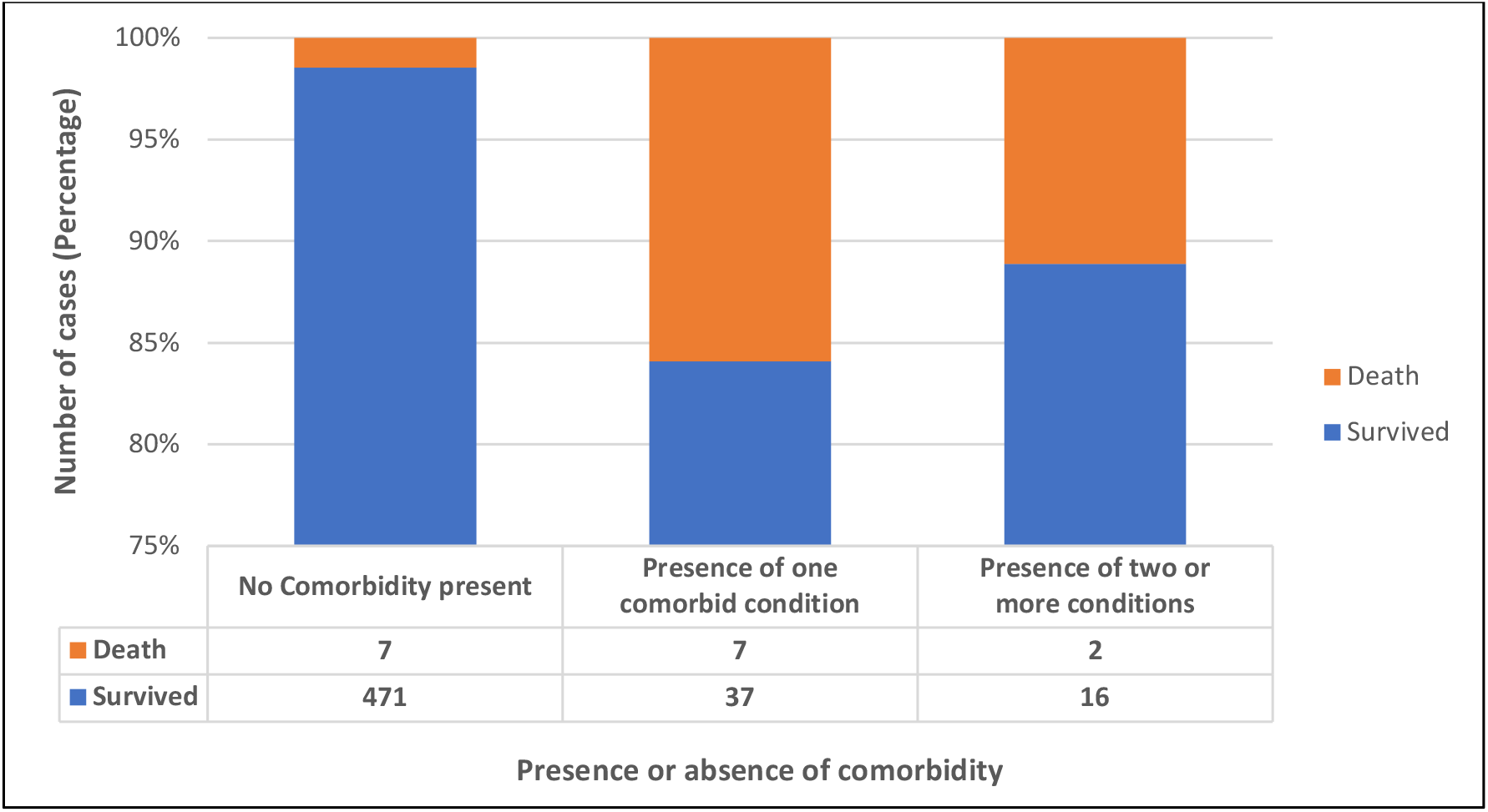
Presence or absence of comorbidity versus the outcome of the disease

**Figure 4:**
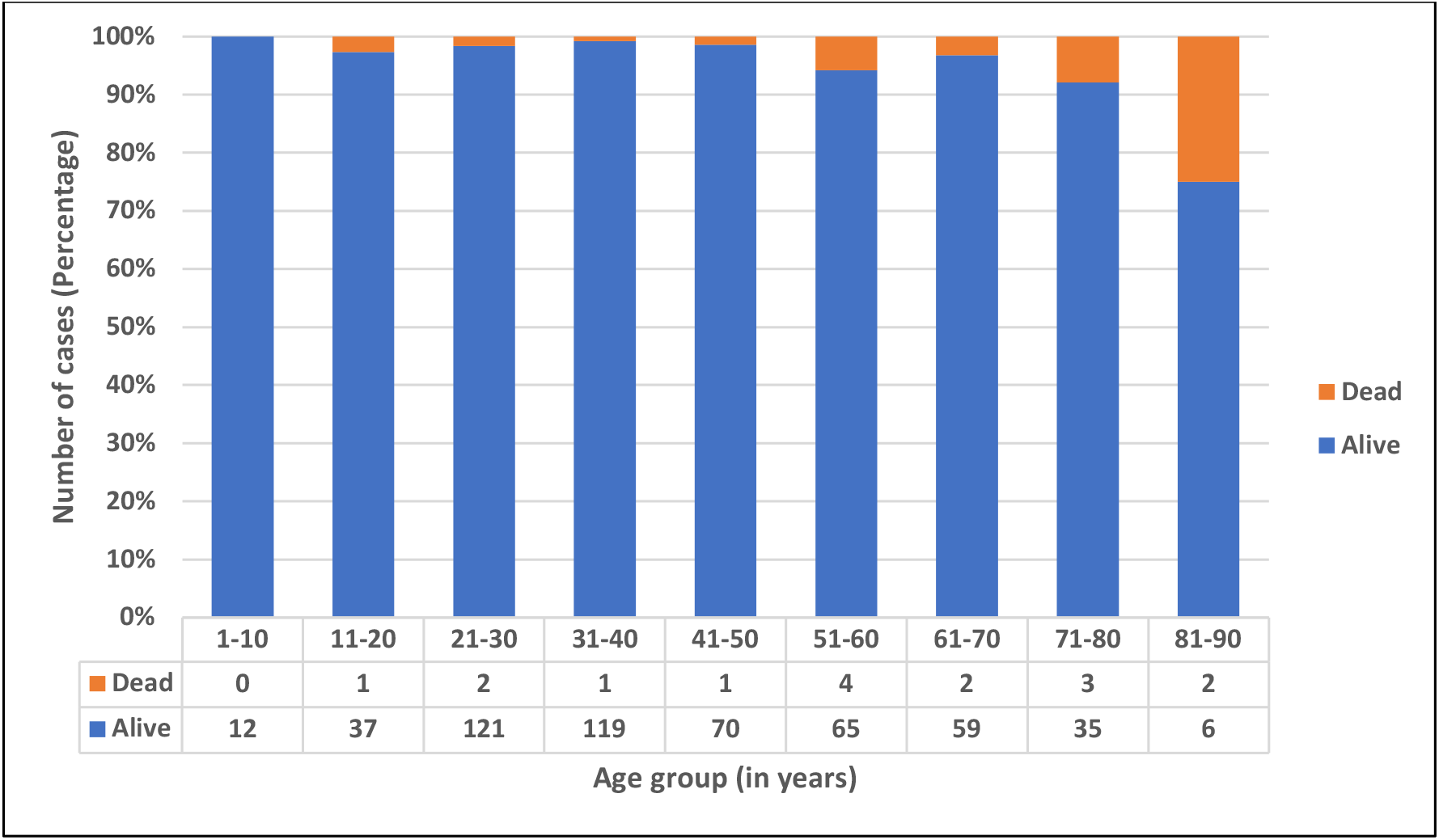
Age-wise distribution of survived and dead cases

Among the 540 cases, 491 (90.93%) were vaccinated with at least a single dose of the COVID-19 vaccine, 41 (7.59%) were unvaccinated, and for eight (1.48%) cases, vaccine data was not available. ***Figure 5*** enumerates the type of vaccine administered to the study population, with COVISHIELD™ (ChAdOx1 nCoV-19 Corona Virus Vaccine) (80.04%) being the most common vaccine, followed by COVAXIN® (BBV152A-a whole inactivated virus-based COVID-19 vaccine) (11.20%). ***Figure 6*** describes the vaccination status of the study population for the age groups. Most unvaccinated individuals fall in the age group of 0 to 10 years (25%) who were not offered vaccination as a part of the vaccination policy in the country. ***Figure 7*** shows the impact of vaccination on the survival of cases. Out of 532 cases with available vaccination data, 491 (92.29%) were vaccinated with at least one dose of vaccine, of which 479 (97.56%) survived, and 12 (2.44%) died. Similarly, out of 41 (7.71%) cases who were not vaccinated, 37 (90.24%) survived, and 04 died (9.76%). The impact of vaccination on the disease outcome was statistically significant, with *p*-value= 0.001.

**Figure 5:**
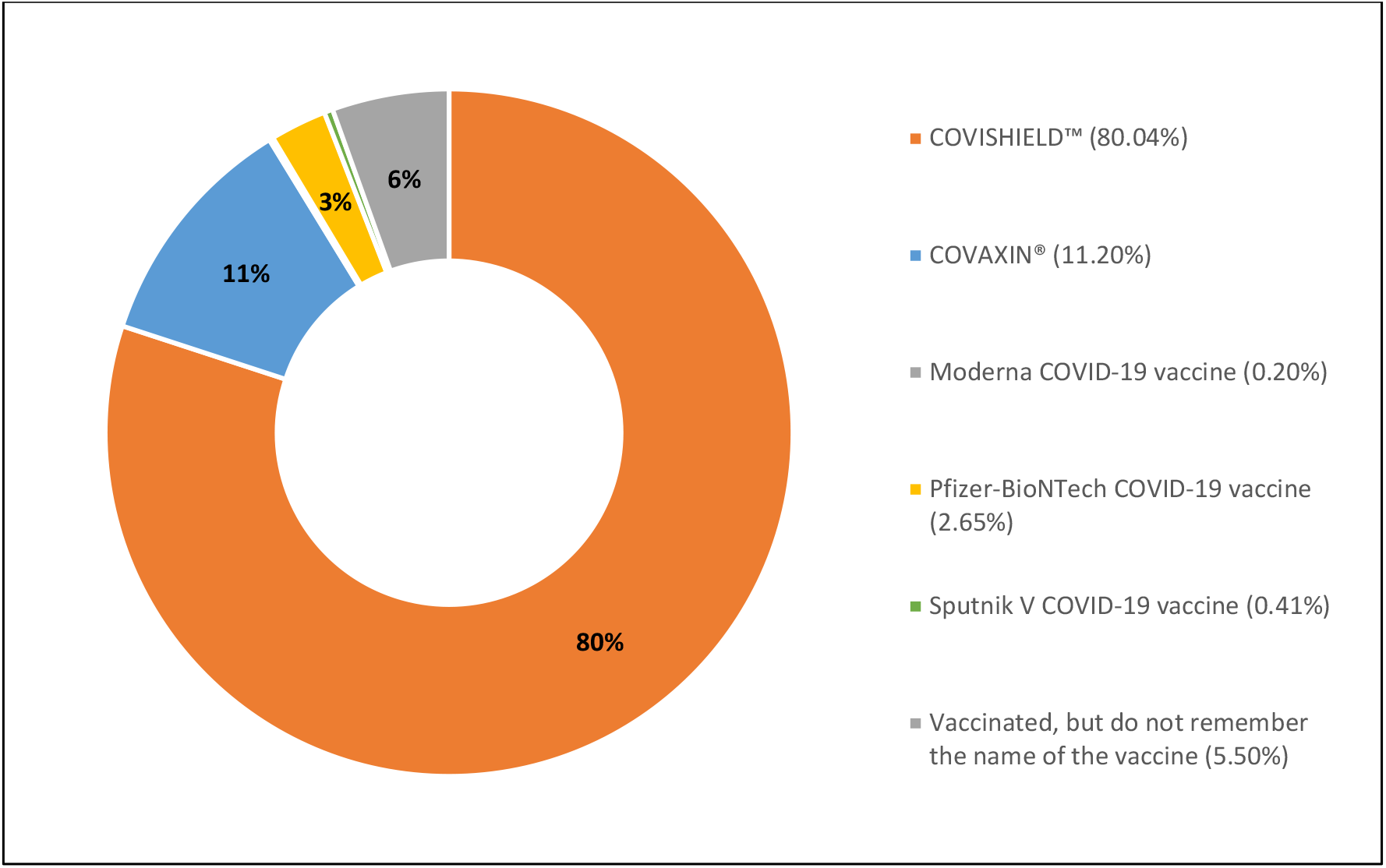
Type of vaccine administered to the study population

**Figure 6:**
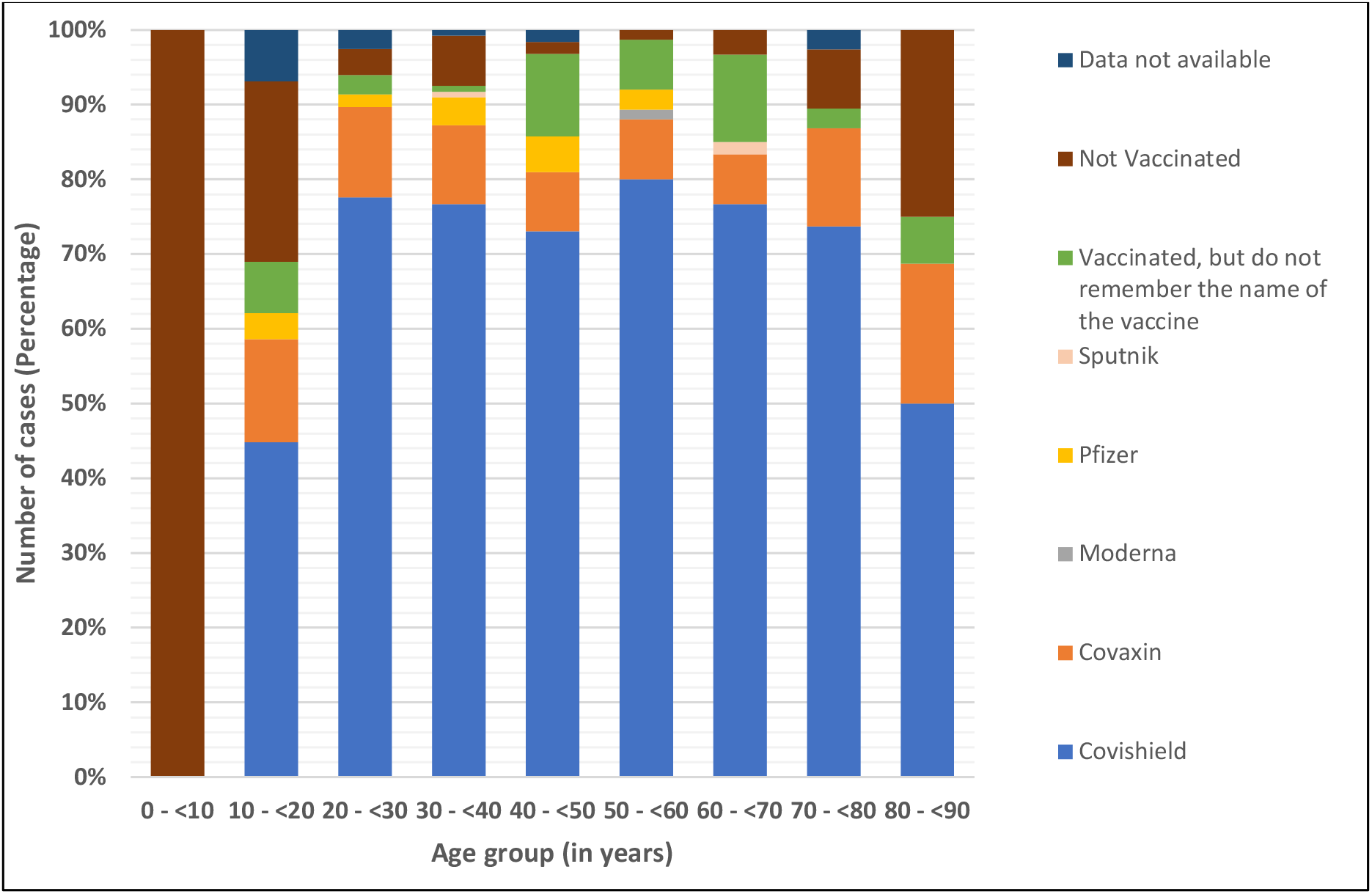
PVaccination status by age groups

**Figure 7:**
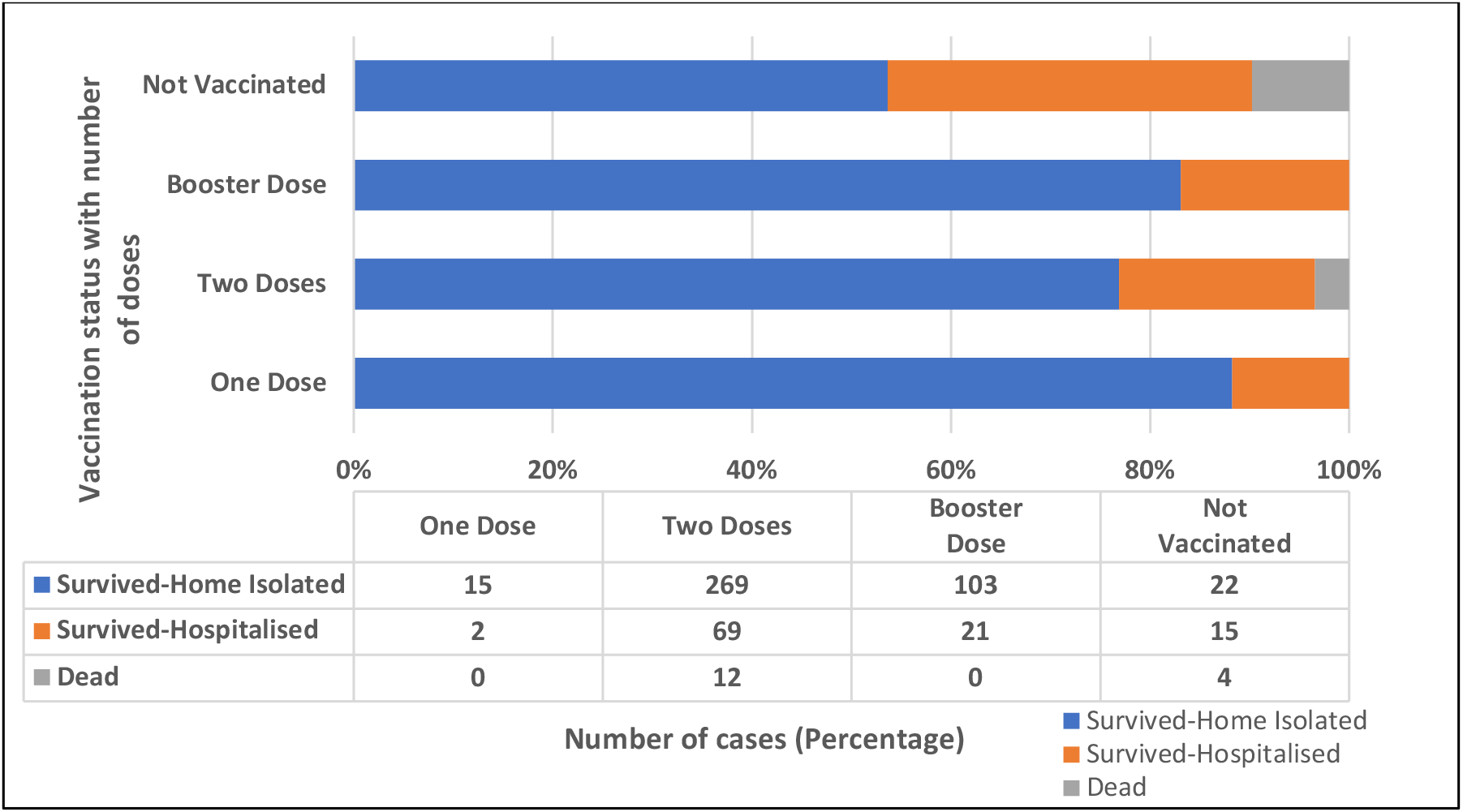
Impact of vaccination on the survival of cases

### 3.4 Clinical characteristics of XBB.1.5 SARS-CoV-2 variant detected during the study

Of the seven XBB.1.5 cases, history was available for six cases. Three out of six cases (50%) had a history of previous infection. All six cases had mild symptoms, with cold and rhinorrhoea (100%) being the most common symptom, followed by sore throat (50%), fever (66.67%) and cough (33.33%). None required hospitalisation or supplemental oxygen; all recovered with conservative treatment at home. Out of the six cases, four had a history of international travel to the United States of America (USA) and Germany, one to Gujarat, and one had contact with a positive person with a history of travel to the USA. All six cases were vaccinated with at least one dose of vaccine, of which 02 (33.33%) cases had received two doses, and 04 (66.67%) had received booster doses. No death was reported among the detected XBB.1.5 cases.

## 4. DISCUSSION

The SARS-CoV-2 virus is known for its unique evolutionary characteristic compared to other respiratory viruses (***Figure 6***). In 2022, saltation or ‘variant’ evolution gave rise to “second generation variants” evolving from a BA.2 lineage background that have numerous non-synonymous mutations, concentrated in the N-terminal domain (NTD) and the receptor binding domain (RBD) of the Spike protein. A few examples include BA.2.75, BA.2.10.4, BJ.1, BS.1, BA.2.3.20, BA.2.83, BP.1, and DD.1, of which BA.2.75 is the most widespread second-generation BA.2 variant. (9) Unlike previous dominant lineages, BA.5 accumulated potent antigenic mutations in a step-wise manner as a result of antigenic drift. The most rapidly growing sublineage of BA.5 is BQ.1, of which BQ.1.1 is the largest containing three further antigenic mutations. (9) SARS-CoV-2 virus, being a coronavirus, is prone to inter-lineage recombination. It generally occurs when a wave declines and a new variant emerges. (10) The resultant recombinant variant possesses unique advantageous properties from both parents. (11) There are 62 recombinant lineages designated by Pangolin (as of December 2022) (12), denoted by a prefix X, of which XBB is the most widespread inter-lineage recombinant to date. (9) XBB is a recombination of BJ.1 (5’ part of XBB genome) and BM.1.1.1 (3’ part of XBB genome) with a breakpoint between 22,897 and 22941 positions in the RBD of the spike protein (corresponding to amino acid positions 445-460). (9, 5) XBB.1.5, a rapidly growing subvariant of the XBB.1 variant, carries an additional mutation F486P in its spike protein, a rare two-nucleotide substitution compared to its parent strain. (13) ***Figure 7*** depicts the mutation prevalence across lineages BJ.1 and BM.1.1.1 and XBB* variant. (14) XBB is shown to have substantially higher viral fitness (Re) than its parental lineages, making it the first documented example of a SARS-CoV-2 variant with increased fitness through recombination. (15)

**Figure 6:**
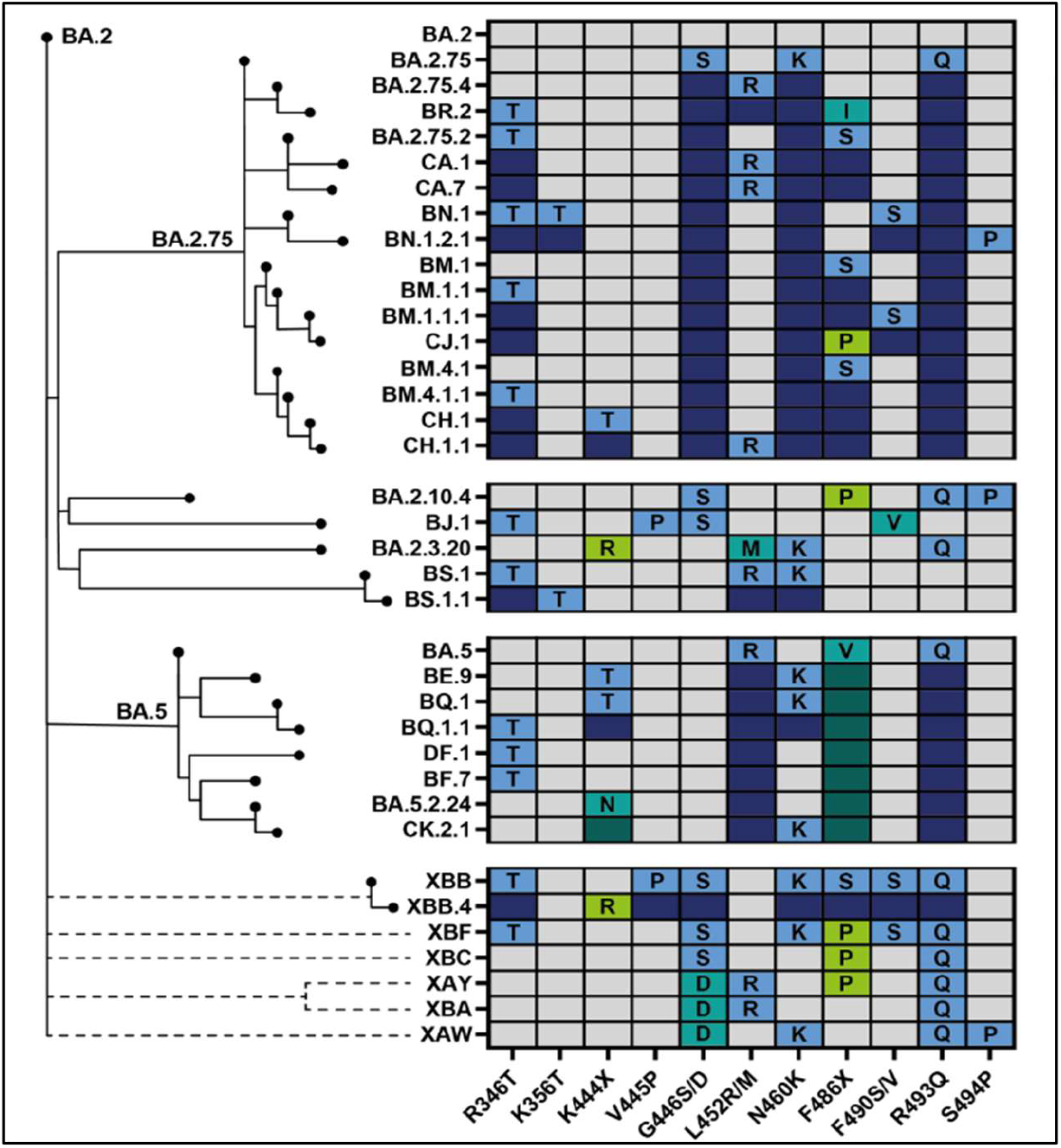
Phylogenetic relatedness and convergent evolution of newer SARS-CoV-2 lineages (9)

**Figure 7:**
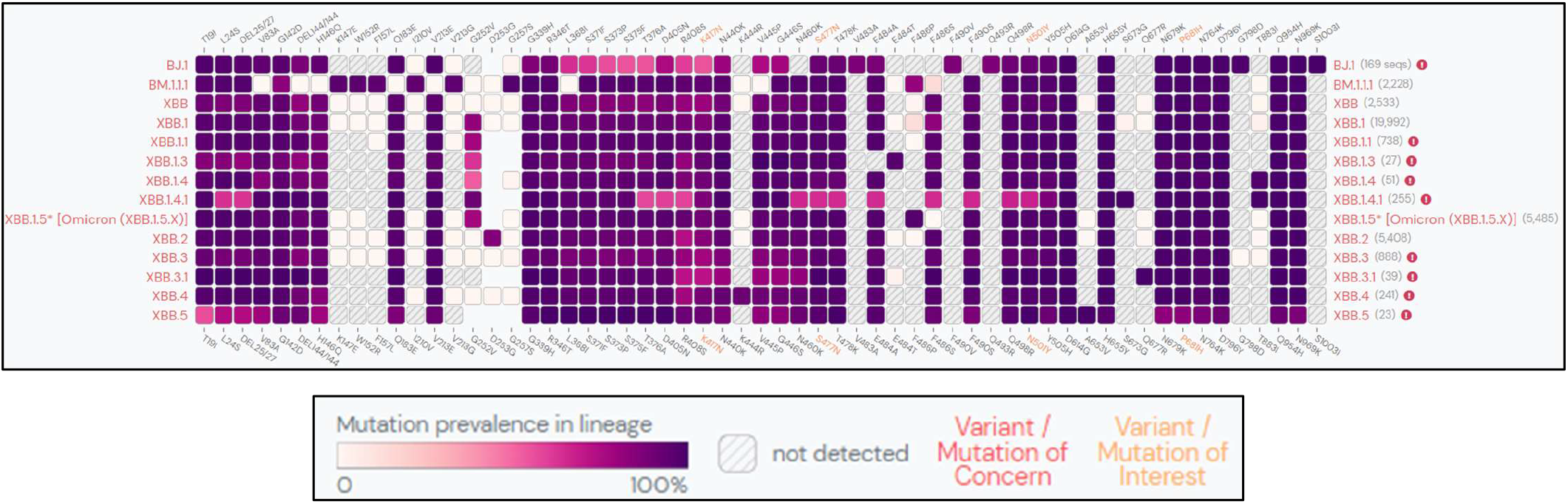
Mutation prevalence across lineages BJ.1, BM.1.1.1 and XBB* (13) (Mutations with > 75% prevalence in at least one lineage)

In the present study, between 10th July 2022 and 12th January 2023, BA.2.75* was the predominant Omicron variant. In India, as of 24^th^ January 2023, the apparent cumulative prevalence of BA.2.75*, XBB*, BA.2.38*, BA.2.10*, BA.5*, BQ.1 * and XBB.1.5 is 14%, 15.68%, 7%, 17%, 4% and 3%, respectively. (16, 17, 18-20, 4, 7) Over the last 60 days, XBB.1 is the dominant lineage (17%) in the country (***Figure 8***). (21) XBB has also spread to countries like Oman, Dominican Republic, Malaysia, Iraq, Singapore, Indonesia and Uganda, with a prevalence of 25% to 93% in the last 60 days as on 24^th^ January 2023. (6) On the contrary, BA.5 and its descendant lineages dominate globally, with about 70.5% of sequences submitted to GISAID between 26^th^ December 2022 to 1^st^ January 2023. (2) However, the prevalence of BA.5 is decreasing globally with a rise in BA.2 descendant lineages, particularly BA.2.75*. The worldwide apparent cumulative prevalence of BQ.1*, BA.5*, BA.2.75*, XBB * and XBB.1.5 is 2%, 14%, 1%, 2.48% and 1%, respectively (as of 24^th^ January 2023). (4, 20, 16, 17, 7) The regional difference in the prevalence of XBB and BQ.1 lineages, XBB being more dominant in the eastern hemisphere and BQ.1 in the western hemisphere, may be due to the proximity of these regions to places where these lineages originated. (15) However, the recently detected XBB.1.5 variant is shown to have growth advantage over other circulating Omicron sublineages, and has already been reported from 46 countries. (7) As of 24^th^ January 2023, a total 9,173 sequences have been deposited on GISAID with the majority of sequences from the USA (74.63%), United Kingdom (9.55%), Canada (3.02%), Austria (2.57%) and Denmark (2.02%). (22)

**Figure 8:**
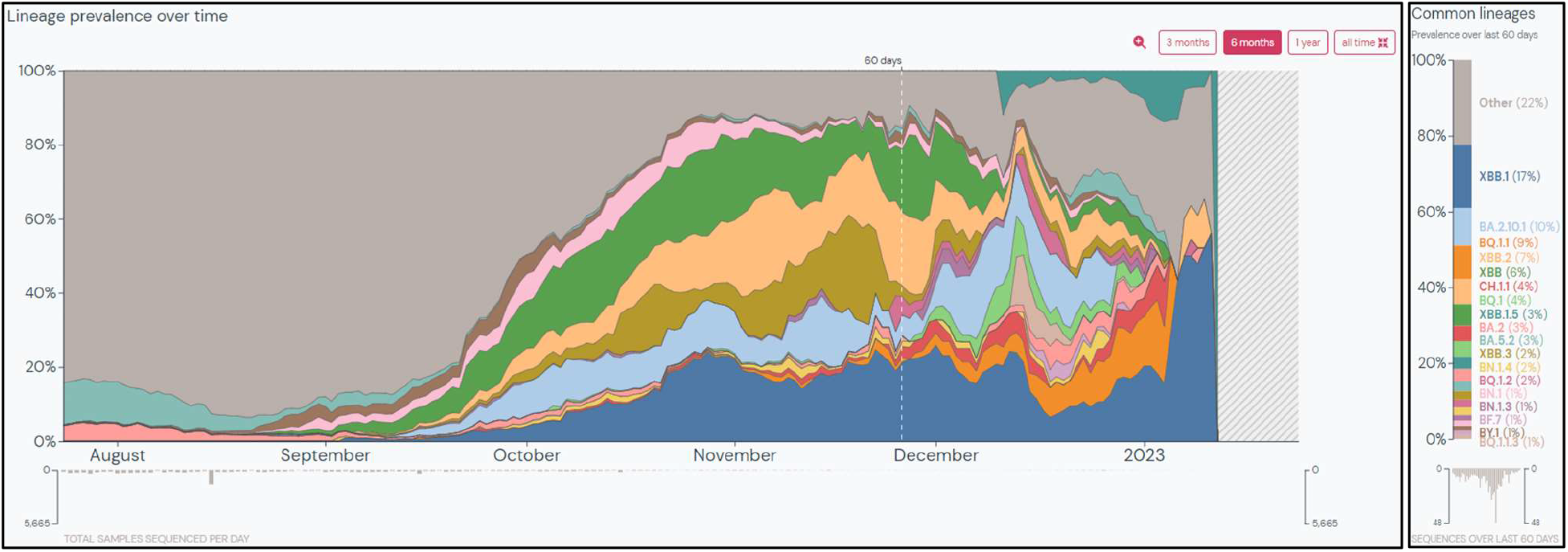
Prevalence of common lineages in India over the last 60 days (as of 24^th^ January 2023) (21)

The most remarkable feature of XBB is its profound resistance to humoral immunity induced by infections with prior Omicron variants. In-vitro studies have shown that XBB exhibited 30-fold and 13-fold resistance to BA.2 and BA.5 infection sera. (15) Similar findings were reported by Wang Q et al., where XBB was ∼63-fold and ∼49-fold more resistant to neutralisation than BA.2 and BA.4/5, respectively. (5) Several NTD and RBD spike mutations like V83A, Y144del, Q183E, R346T, L368I, V445P, F486S and F490S cooperatively contribute to resistance against humoral immunity induced by BA.2 infections. Similarly, two mutations, Y144del and G446S, were suggested to contribute to resistance against humoral immunity induced by BA.5 infections. The humoral immune-escape property of XBB.1.5 was comparable to XBB.1. (13) Spike mutations in the RBD (R346T, L368I and N460K) and NTD (V83A) are also responsible for increased ACE2 binding affinity, viral infectivity and fusogenicity of XBB compared to BA.2, BA.5 and BA.2.75 Omicron variants. (15) Deep mutational scanning studies have shown that due to the presence of additional Ser486Pro mutation in the spike protein, XBB.1.5 had a stronger binding affinity to ACE2 receptors than XBB.1, thereby explaining its significant growth advantage over XBB.1. (13) XBB-infected hamster sera showed a remarkable antiviral effect against XBB only, suggesting XBB is antigenically distinct from other Omicron subvariants. (15)

Several clinically authorised therapeutic monoclonal antibodies (mAbs) like bamlanivimab, etesevimab, imdevimab, casirivimab, tixagevimab, cilgavimab, and sotrovimab have been rendered ineffective by previous SARS-CoV-2 variants, leaving bebtelovimab as the only monoclonal antibody active against the circulating strains. However, due to mutations like N460K, F486S, R346T, V455P, G446S and F490S, XBB and its descendant XBB.1 are pan-resistant to RBD class I, II and III antibodies. (5) Also, bebtelovimab (LY-CoV1404) and Evusheld (a combination of COV2-2196 and COV2-2130) were found to be inactive against XBB/XBB.1 (5) and XBB.1.5 (13). Although XBB.1.5 escapes neutralising antibody responses, a study by Lasrado N et al. has shown that the cross-reactive T-cell responses may continue to protect against severe disease. (23) Therefore, due to immune-escape-associated and infectivity-enhancing mutations, the XBB* and XBB.1.5 variants can eventually spread worldwide. (15)

The present study suggests that the pathogenicity of XBB is comparable to that of other Omicron variants. These findings are consistent with an in-vivo study in hamsters where the XBB variant was less pathogenic than the Delta variant and had a comparable pulmonary function and viral RNA load to BA.2.75 infected hamsters. The intrinsic pathogenicity of the XBB variant and its efficiency of infecting lungs was comparable to or even lower than the BA.2.75. (15) Similarly, the seven XBB.1.5 cases detected had mild symptoms. However, it remains unclear whether the intrinsic pathogenicity of the virus or the immunity from vaccination and previous infection is responsible for mild cases in India. More clinical data from other clinical settings with different levels of immunity will help understand the behaviour of the XBB* variant.

## 5. CONCLUSION

To conclude, while it is encouraging that the infection caused by the SARS-CoV-2 XBB recombinant variant tends to be less severe and similar to that of other Omicron subvariants, the fact that this variant is more transmissible and immune-evasive is a cause of concern. In addition to the ongoing surveillance efforts to track the spread of new variants, our findings emphasise the importance and the need to monitor the clinical severity of infections caused by new variants. Such findings are essential for making decisions about deploying interventions and preparing healthcare systems to respond to outbreaks.

## Data Availability

All data produced in the present work are contained in the manuscript.

## ACKNOWLEDGEMENT

We thank Mrs Poonam Pacharne, Mr Vishal Rajput and Mrs Reena Katke from Byramjee Jeejeebhoy Government Medical College and Sassoon General Hospitals, Pune; Miss Aditi and Miss Pratiksha from ICMR-National Institute of Virology, Pune, for their technical help during the study.

